# A rapid review of the effectiveness of alternative education delivery strategies in medical, dental, nursing and pharmacy education during the COVID-19 pandemic

**DOI:** 10.1101/2022.03.04.22271892

**Authors:** Judith Carrier, Deborah Edwards, Michal Tombs, Steve Riley, Ruth Lewis, Elizabeth Gillen, Alison Cooper, Adrian Edwards

## Abstract

**Background:** Education delivery in higher education institutions was severely affected by the COVID-19 pandemic, with emergency remote teaching developed and adapted promptly for the circumstances. This rapid review investigated the effectiveness of alternative education delivery strategies during the pandemic for medical, dental, nursing and pharmacy students, to help plan and adapt further education provision.

**Methods:** We included 23 primary studies in undergraduate education, all published in 2020-2021, no relevant UK-based or postgraduate studies were found. Included studies comprised 10 single cohort descriptive; 11 comparative descriptive; and two RCTs. There was considerable variability in terms of students, type of distance learning, platforms used and outcome measures.

**Results:** In medicine (n=14), self-reported competency and confidence, and demonstrable suturing skills were achieved through participating in remote learning. However, lower levels of knowledge were obtained by students who received virtual or blended learning compared to in-person teaching (low-very low confidence). Using bespoke interactive platforms in undergraduate medical training was superior to standard video (low confidence) or ‘textbook’ presentations (very low confidence).

In dentistry (n=2), remote learning led to knowledge gained (low confidence), but self-reported practical and interpersonal skills were lower with remote rather than in-person learning (very low confidence).

In nursing (n=3), remote learning, when compared to in-person, resulted in similar knowledge and self-reported competency levels (very low confidence) pre-COVID, but confidence was higher when learning or assessment was conducted virtually (low confidence).

In pharmacy (n=4), virtual learning was associated with higher skills, but lower knowledge compared to in-person, pre-COVID; self-reported competency and confidence scores were similar between the two groups (very low confidence).

**Conclusions:** Remote teaching was valued, and learning was achieved, but the comparative effectiveness of virtual versus in-person teaching is less clear. Supplementary alternative or in-person practical sessions may be required post-emergency to address learning needs for some disadvantaged student groups.

## Introduction

Education delivery in higher education institutions was severely affected by the COVID-19 pandemic, especially for healthcare students whose continuing education is imperative to maintain a well-educated healthcare workforce. Many courses transitioned to a period of remote emergency teaching,^1–3^ developed and adapted promptly for the circumstances, largely without prior contingency planning. For example the American Society of Plastic Surgeons announced free access to its online Education Network for all medical students with an interest in plastic surgery,^4^ whilst Ahmed at al^5^ suggested a range of online tools and resources that could be employed for online rheumatology education. In Jordan, distance e-learning was promptly engaged to maintain the continuity of medical education,^6^ and in Pakistan dental educators came up with innovative solutions to resume dental education remotely.^7^ Bakshi et al^8^ argued that whilst the COVID-19 outbreak disrupted the educational experiences of medical students worldwide, this was particularly significant in areas such as ophthalmology where structured education and clinical exposure had already declined. A shift to virtual education for nursing students in Iran highlighted some of the challenges faced by educators and students, such as lack of infrastructure, reduced readiness of educators and students for e-learning, and the time to prepare educational content,^9^ whilst educators in Canada^10^ emphasised the importance of continuing to engage nursing students online. Reviews have also highlighted the challenges in migrating to remote education^11,12^ which include poor knowledge of staff on how to deal with technology, poor internet connections and difficulty in transitioning content for online learning.^11,12^ By contrast, some students and staff report satisfaction with remote learning,^2,13^ especially when collaboration and engagement with peers is facilitated.^2^

A preliminary search of repositories specific to COVID-19 literature identified several existing reviews of alternative education delivery strategies for medical and healthcare students during the COVID-19 pandemic. The systematic review by Wilcha et al.^3^ looked at the effectiveness of virtual teaching for medical education and suggested that it was effective. However, searching was limited to two databases, including Google Scholar, and the review appears to have been conducted by one author with no critical appraisal conducted.^3^ Another systematic review by He et al.^13^ explored the use of synchronous distance education (videoconference or web conference, online classroom or virtual classroom) compared with traditional education for medical, dental, nurse, pharmacy students and other health science–related students.^13^ It found that there were no significant differences in terms of knowledge or skills but that satisfaction was rated higher for distance education.^13^ For nursing students, a scoping review by Jowsey et al.^14^ suggested that when delivered purposefully, blended learning (a mix of face-to-face and online study) can positively influence and impact on the achievements of students, especially when used to support distance education.^14^ However, none of the existing reviews specifically explored effectiveness of alternative education delivery strategies for medical, nursing, dental and pharmacy students, or allied health professionals during the COVID-19 pandemic, or provided a separate summary of the evidence for these disciplines. An initial scope of the evidence base for these healthcare disciplines identified a significant volume of primary research in the area for medical, nursing, dental and pharmacy students but very little for other healthcare disciplines including allied health professionals. We therefore conducted a rapid review of the effectiveness of alternative education delivery strategies that have been put into place for undergraduate and postgraduate medical, nursing, dental, and pharmacy students.

## Methods

This rapid review was registered with the International prospective register of systematic reviews (PROSPERO) following the completion of the database searches, and study selection (Registration number: CRD42022304295).

### Eligibility criteria

The inclusion criteria were informed by the PICO (Participants, Intervention, Comparison, Outcomes) framework (see Table 1).

**Table 1:** Eligibility criteria.

| PICO | Inclusion criteria | Exclusion criteria |
| --- | --- | --- |
| Participants | Undergraduate students<br>Post-graduate students<br>Medicine, Dentistry,<br>Nursing, Pharmacy | All other allied health professions |
| Intervention / exposure | Specific educational delivery (including clinical skills delivery) during COVID-19 | Assessment / examination processes<br>Continuing professional development not leading to a postgraduate qualification |
| Comparison | Education delivery (including clinical skills delivery) prior to COVID-19 |  |
| Outcomes | Educational outcomes of knowledge, skills, confidence, competency |  |
| Further study considerations |  |  |
| Study design | Primary research |  |
| Context | All academic and healthcare institutions that deliver undergraduate or post graduate education with OECD countries |  |

### Search strategy

Searches were conducted across four databases: On the OVID platform: MEDLINE and Embase, on the EBSCO platform: CINAHL and ERIC, from December 2019 to 8th June 2021 for English language citations. An initial search of MEDLINE was undertaken (medicine or medical or nurs* or dental or dentistry or pharmacy or pharmacist) AND education* or train* or teach* or student* or undergraduate* or postgraduate* AND COVID* or coronavirus) followed by analysis of the text words contained in the title and abstract, and of the index terms used to describe the article. This informed the development of search strategies tailored for each information source (additional material one). The reference list of all included studies was screened for additional studies.

### Study selection process

All citations retrieved from the database searches were imported into EndNote™ and duplicates and irrelevant citations removed and then imported to Covidence™ for study selection. Two reviewers dual screened at least 20% of citations using the information provided in the title and abstract using the software package Covidence™, resolving all conflicts. The remaining citations were then screened by a single reviewer, screening with categories of ‘include’ and ‘exclude’. To streamline the review process, the project team decided against a third category of ‘unsure’ and instead, where there was uncertainty about a citation, it was categorised as ‘include’ and the decision was made based on the full text. The full texts were screened for inclusion by one reviewer using a purposefully designed form which was piloted using approximately 10 manuscripts. One reviewer then screened full text manuscripts, and another reviewer checked all excluded manuscripts.

### Data extraction

All demographic data were extracted directly into tables by one reviewer and checked by another. The data included specific details about the interventions, populations, study methods and outcomes of significance to the review question and specific objectives. A template for the data extraction process was piloted on manuscripts for each of the included study designs before use. All outcome data were extracted directly into tables by one reviewer and checked by another.

### Quality appraisal

The methodological quality of all the research studies was assessed by one reviewer, and judgements verified by a second reviewer, using JBI design-specific critical appraisal tools (https://jbi.global/critical-appraisal-tools). When a study met a criterion for inclusion a score of one was given. Where a particular point for inclusion was regarded as “unclear” it was given a score of zero. Where a particular point for inclusion was regarded as “not applicable” this point was deducted from the total score. All included studies were assessed using this method and their overall critical appraisal scores were calculated and are displayed for each study in Tables 2 and 3. For the full details of the critical appraisal scores see additional material two.

**Table 2:**
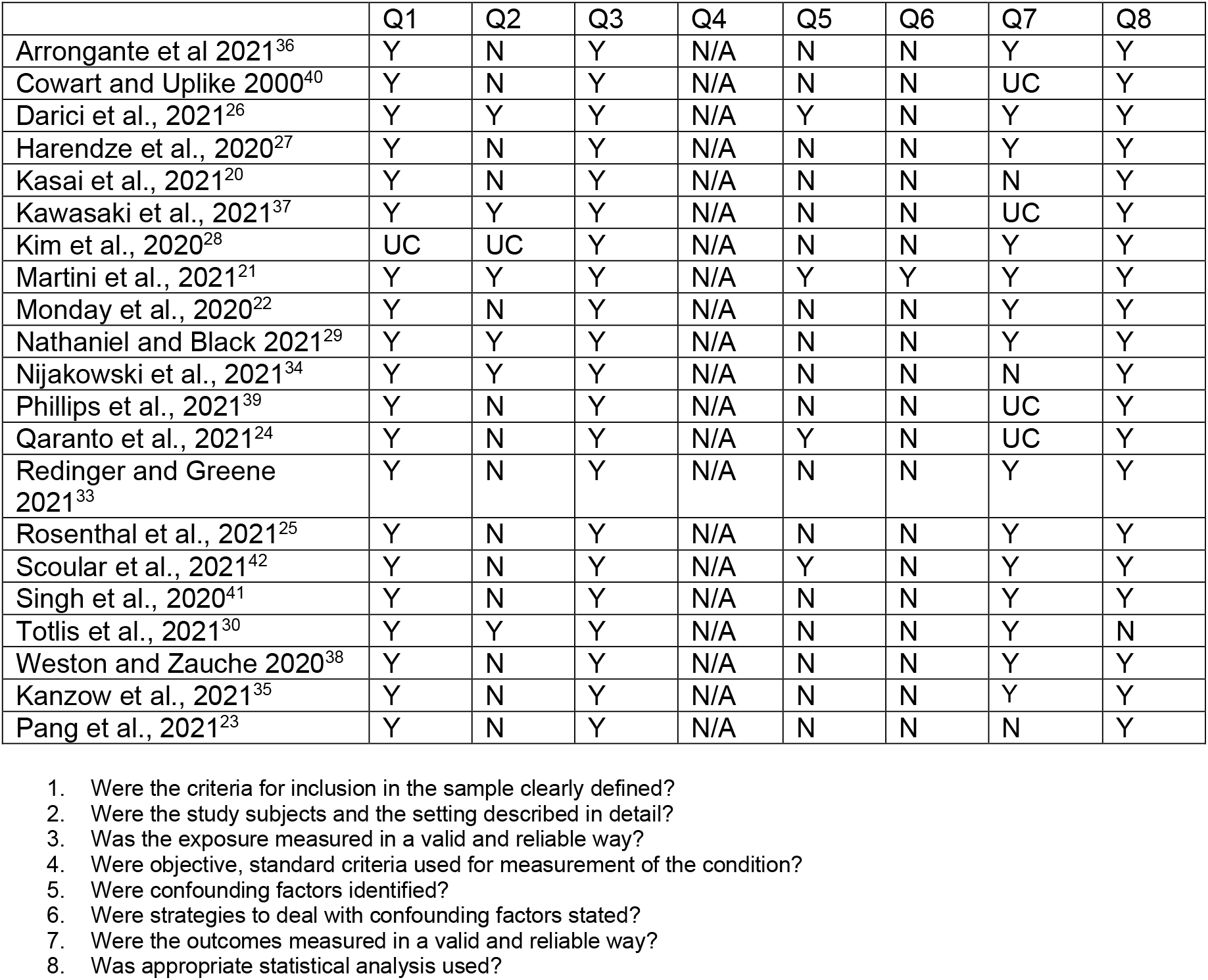
Summary of critical appraisal scores from descriptive surveys.

|  | Q1 | Q2 | Q3 | Q4 | Q5 | Q6 | Q7 | Q8 |
| --- | --- | --- | --- | --- | --- | --- | --- | --- |
| Arrongante et al 2021<sup>36</sup> | Y | N | Y | N/A | N | N | Y | Y |
| Cowart and Uplike 2000<sup>40</sup> | Y | N | Y | N/A | N | N | UC | Y |
| Darici et al., 2021<sup>26</sup> | Y | Y | Y | N/A | Y | N | Y | Y |
| Harendze et al., 2020<sup>27</sup> | Y | N | Y | N/A | N | N | Y | Y |
| Kasai et al., 2021<sup>20</sup> | Y | N | Y | N/A | N | N | N | Y |
| Kawasaki et al., 2021<sup>37</sup> | Y | Y | Y | N/A | N | N | UC | Y |
| Kim et al., 2020<sup>28</sup> | UC | UC | Y | N/A | N | N | Y | Y |
| Martini et al., 2021<sup>21</sup> | Y | Y | Y | N/A | Y | Y | Y | Y |
| Monday et al., 2020<sup>22</sup> | Y | N | Y | N/A | N | N | Y | Y |
| Nathaniel and Black 2021<sup>29</sup> | Y | Y | Y | N/A | N | N | Y | Y |
| Nijakowski et al., 2021<sup>34</sup> | Y | Y | Y | N/A | N | N | N | Y |
| Phillips et al., 2021<sup>39</sup> | Y | N | Y | N/A | N | N | UC | Y |
| Qaranto et al., 2021<sup>24</sup> | Y | N | Y | N/A | Y | N | UC | Y |
| Redinger and Greene 2021<sup>33</sup> | Y | N | Y | N/A | N | N | Y | Y |
| Rosenthal et al., 2021<sup>25</sup> | Y | N | Y | N/A | N | N | Y | Y |
| Scoular et al., 2021<sup>42</sup> | Y | N | Y | N/A | Y | N | Y | Y |
| Singh et al., 2020<sup>41</sup> | Y | N | Y | N/A | N | N | Y | Y |
| Totlis et al., 2021<sup>30</sup> | Y | Y | Y | N/A | N | N | Y | N |
| Weston and Zauche 2020<sup>38</sup> | Y | N | Y | N/A | N | N | Y | Y |
| Kanzow et al., 2021<sup>35</sup> | Y | N | Y | N/A | N | N | Y | Y |
| Pang et al., 2021<sup>23</sup> | Y | N | Y | N/A | N | N | N | Y |

1. Were the criteria for inclusion in the sample clearly defined? 2. Were the study subjects and the setting described in detail? 3. Was the exposure measured in a valid and reliable way? 4. Were objective, standard criteria used for measurement of the condition? 5. Were confounding factors identified? 6. Were strategies to deal with confounding factors stated? 7. Were the outcomes measured in a valid and reliable way? 8. Was appropriate statistical analysis used?

**Table 3:** Summary of critical appraisal scores from randomised controlled trials.

|  | Q1 | Q2 | Q3 | Q4 | Q5 | Q6 | Q7 | Q8 | Q9 | Q10 | Q11 | Q12 | Q13 |
| --- | --- | --- | --- | --- | --- | --- | --- | --- | --- | --- | --- | --- | --- |
| Suppan et al., 2021<sup>32</sup> | Y | Y | Y | N/A | N/A | Y | Y | N | Y | Y | Y | Y | Y |
| Schmitz et al., 2021<sup>31</sup> | Y | Y | UC | N/A | N/A | UC | Y | N | Y | Y | Y | Y | UC |

1. Was true randomization used for assignment of participants to treatment groups? 2. Was allocation to treatment groups concealed? 3. Were treatment groups similar at the baseline? 4. Were participants blind to treatment assignment? 5. Were those delivering treatment blind to treatment assignment? 6. Were outcomes assessors blind to treatment assignment? 7. Were treatment groups treated identically other than the intervention of interest? 8. Was follow up complete and if not, were differences between groups in terms of their follow up adequately described and analysed? 9. Were participants analysed in the groups to which they were randomized? 10. Were outcomes measured in the same way for treatment groups? 11. Were outcomes measured in a reliable way? 12. Was appropriate statistical analysis used? 13. Was the trial design appropriate, and any deviations from the standard RCT design (individual randomization, parallel groups) accounted for in the conduct and analysis of the trial?

### Synthesis

The data were reported narratively as a series of thematic summaries^15^ and presented separately for each health care discipline. Two RCTs were included in the review but there was insufficient homogeneity across the studies and therefore we were unable to perform a meta-analysis.

### Assessment of body of evidence

The confidence in the synthesised findings was assessed by one reviewer and judgements verified by a second reviewer. The RCTs were assessed using the Grading of Recommendations, Assessment, Development and Evaluation (GRADE) approach.^16^ Due to heterogeneity of the different interventions within similar settings, outcome data were only available for results from single studies and guidance was followed on undertaking GRADE for data of this type.^17^ Quantitative descriptive studies were assessed by applying the principles of GRADE.^18^ For further details of this processes see additional material three and four. Most findings in this rapid review were of low or very low quality and ratings are displayed for each study in Tables 2 and 3. This was mainly due to imprecision because of small sample sizes, and/or confidence intervals not being reported, and/or limitations because baseline levels of the outcome of interest not being controlled for, and/or lack of clarity of confounding factors.

## Results

Of the 10,978 citations retrieved from our searches, 21 descriptive studies and two RCTs met our eligibility criteria. For details of the excluded studies see additional material five. The included studies focused on undergraduate medical students (n=14), undergraduate dental students (n=2), undergraduate nursing students (n=3) and undergraduate pharmacy students (n=4). We did not find any studies that focused on postgraduate students, and research, that focused on clinically based postgraduate training, such as internships, were excluded. The flow of citations through each stage of the review process is displayed in a PRISMA flowchart, ^19^ see Figure 1.

**Figure 1:**
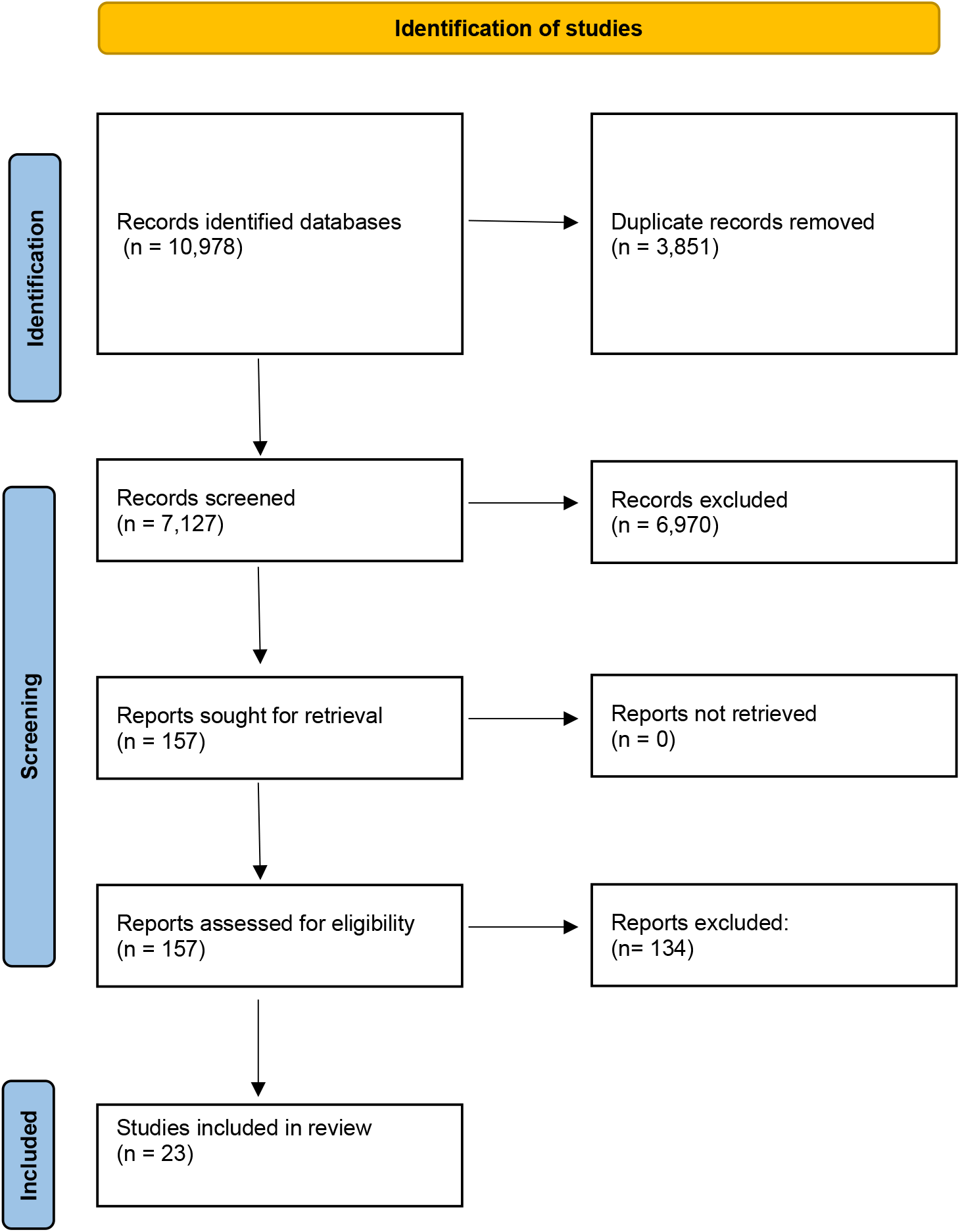
PRISMA flow diagram

### Overview of evidence base for medical students

Six pre-test / post-test designs^20–25^ and six post-test only descriptive studies.^26–30^ and two RCTs,^31,32^ provided evidence of the effectiveness of alternative education delivery strategies for undergraduate medical students during the COVID-19 pandemic (see Table 4). Most studies (n=7) were conducted in the USA.^21–25,29,33^ The remaining studies were conducted in Germany,^26,27,31^ Japan,^20^ South Korea,^28^ Switzerland^32^ and Greece.^30^

**Table 4:** Characteristics of included studies focusing on medical students.

| Author/s<br>Country | Participants | Study design<br>Type of analysis | Findings |
| --- | --- | --- | --- |
| Focus<br>Remote platform | Outcomes/outcome measures |  |  |
| Martini et al., 2021<sup>21</sup><br>USA<br>Virtual neurosurgery seminar series<br>Zoom video conferencing platform<br>16 one-hour seminars that were conducted biweekly over the course of a 2-month period | <u>Participants</u><br>June, July 2020<br>595 medical students (from all school years 1 to 5) across the countries registered with an average of 82 students participating live in each weekly lecture (range, 41-150)<br>Completing pre and post-test study (n=32)<br><u>Outcomes</u><br>Confidence with material pertaining to core concepts across various neurosurgical subdisciplines.<br><u>Outcome measures</u><br>Self-assessment scale of 1-10 (1=not confident at all; 10= very confident) | <u>Study design</u><br>Descriptive study<br>Pre-test / Post-test<br><u>Type of analysis</u><br>Analytical statistics<br>Mean scores<br><u>Quality appraisal rating</u><br>Score of 6 out of 7<br><u>Confidence evaluation</u><br>Confidence – Low | <u>Confidence (Mean±SD)</u><br>Cerebrovascular neurosurgery<br>Pre (5.90±0.34); Post (8.36±0.19)<br>p<0.0001<br>Malignant brain tumours<br>Pre (4.95± 0.45); Post (8.28 ± 0.27)<br>p<0.0001<br>Head trauma<br>Pre (5.54± 0.34); Post (7.97± 0.27)<br>p<0.0001)<br>Spine trauma<br>Pre (4.96± 0.38); Post (8.19± 0.26)<br>p<0.0001)<br>Neuroendocrinology/pituitary pathology<br>Pre (6.79± 0.31); Post (8.74± 0.19)<br>p<0.0001)<br>Pediatric neurosurgery<br>Pre (5.79± 0.33); Post (8.25±0.26)<br>p<0.0001)<br>Neurocritical care<br>Pre (4.86± 0.44); Post (8.25± 0.26)<br>p<0.0001)<br>Minor neurosurgical procedures<br>Pre (4.48± 0.44); Post (7.86± 0.28)<br>p<0.0001) |
| Monday et al., 2020<sup>22</sup><br>USA<br>Online virtual internship boot camp<br>Residency preparation course<br>Canvas online learning management system<br>26 sessions (22 mandatory and 4 optional) over one month | <u>Participants</u><br>Academic years 2019/2020<br>Fourth years (n=89)<br>Self-assessed confidence and knowledge response rates<br>Pre-test (76–87%)<br>Post-test (60-82%)<br>Post-test assessment<br>Response rate 99%<br><u>Outcomes</u><br>Confidence and knowledge for 14 out of the 26 sessions across the American Academy of Medical Colleges 13 core competencies<br><u>Outcome measures</u><br>5-point self-assessment Likert scale (1 meaning confidence or knowledge was very poor, 3 meaning neutral, and 5 meaning very high) | <u>Study design</u><br>Descriptive study<br>Pre-test / Post-test<br><u>Type of analysis</u><br>Analytical statistics<br>Mean scores<br><u>Quality appraisal rating</u><br>Score of 4 out of 7<br><u>Confidence evaluation</u><br>Confidence – Low<br>Knowledge – Low | <u>Confidence</u><br>A significant increase in self assessed confidence across all the American Academy of Medical Colleges 13 core competencies was demonstrated (p<0.001)<br><u>Knowledge</u><br>A significant increase in self assessed knowledge across all the American Academy of Medical Colleges 13 core competencies was demonstrated (p<0.001)<br>All students passed post-test assessment 83 (94%) achieved a score of 70% or higher, 4 (4.5%) scored in the 60-70% range, and 1 (1.1%) scored 55% |
|  | <u>Knowledge</u><br>53 item competency-based exam |  |  |
| Qaranto et al., 2021<sup>24</sup><br>USA<br>Interactive remote sessions on surgical instruments, knot tying and suturing ("remote coach model")<br>Zoom video conferencing platform<br>Three sessions | <u>Participants</u><br>Academic year 2019/2020<br>Third years enrolled in surgical clerkship (n=31)<br><u>Outcomes</u><br>Knot tying confidence and skills<br>Suturing ability confidence and skills<br><u>Outcome measures</u><br>Visual demonstration of knot tying and suturing<br>Self-assessment of confidence but details of the scale not reported | <u>Study design</u><br>Descriptive study<br>Pre-test / Post-test<br><u>Type of analysis</u><br>Analytical statistics<br>Mean scores<br><u>Quality appraisal rating</u><br>Score of 4 out of 7<br><u>Confidence evaluation</u><br>Confidence – Very low<br>Skills – Very Low | <u>Confidence</u> (Mean±SD)<br>Knot tying<br>Pre (7.86±0.66); Post (9.65±0.85)<br>p=0.028<br>Suturing techniques<br>Pre (8.0±1.3); Post (13.8±0.9),<br>p<0.001<br><u>Skills</u><br>All students successfully demonstrated their ability to tie two handed knots and perform simple sutures |
| Darici et al., 2021; <sup>26</sup><br>Germany<br>Online digital histology course<br>Zoom video conferencing platform<br>19 days | <u>Participants</u><br>Academic year 2019/2020<br>Second years (n=132/192 sat the exam)<br>Third years (n=175/201 sat the exam)<br><u>Outcomes</u><br>Knowledge<br><u>Outcome measures</u><br>Multiple choice final exam | <u>Study design</u><br>Descriptive study<br>Post test<br><u>Type of analysis</u><br>Descriptive statistics<br>% passing exam<br><u>Quality appraisal rating</u><br>Score of 7 out of 7<br><u>Confidence evaluation</u><br>Knowledge –Moderate | <u>Knowledge</u><br>Second years<br>Median was 71% correct answers (SD 18.5%, 95% CI 65%, 72%)<br>Third years including repeating students<br>Median was 74% correct answers (SD 20.2%, CI 67%, 73%)<br>Third years without repeating students<br>Median 76% correct answers (SD 19.8, 95% CI 68%, 75%) |
| Harendza et al., 2020<sup>27</sup><br>Germany<br>Virtual training including simulated patient consultations, documentation, and case presentation<br>Zoom video conferencing platform<br>Training included a consultation hour with four simulated patients per participant, patient documentation and management with a newly developed electronic patient chart, and one case presentation per participant in hand-off format | <u>Participants</u><br>Academic year 2020/2021<br>Final years (n=32)<br>Online learning<br>Academic year 2019/2020<br>Final years (n=103)<br>Clinical learning<br><u>Outcomes</u><br>Confidence<br><u>Outcome measures</u><br>5-point self-assessment Likert scale<br>1=does not apply, 2= somewhat applies, 3=partly applies, 4=rather applies, 5= fully applies | <u>Study design</u><br>Descriptive study<br>Post test<br><u>Type of analysis</u><br>Analytical statistics<br>Mean scores<br>Comparison between remote and in person learning across two academic years<br><u>Quality appraisal rating</u><br>Score of 4 out of 7<br><u>Confidence evaluation</u><br>Confidence – Very low | <u>Confidence</u> (Mean±SD)<br>I felt confident during history taking<br>Clinical learning (3.67±0.87); Virtual (3.88±0.79), p>0.05<br>I felt confident during the management phase time<br>Clinical learning (3.12±0.9); Virtual (3.16±0.72), p>0.05<br>I felt confident during the case presentation<br>Clinical learning (3.33±0.96); Virtual (3.42±0.92), p>0.05 |
| Kasai et al., 2021<sup>20</sup><br>Japan<br>Online simulated clinical practice for the respiratory unit and general medicine<br>Zoom video conferencing platform<br>4 weeks | <u>Participants</u><br>Academic Year 2019/2020<br>Fifth years (Clerkship)(n=43)<br><u>Outcomes</u><br>Competency<br>Across 9 domains<br>Medical interviewing, physical examination, humanistic qualities/professionalism, clinical judgment, counselling, organization or efficiency, overall clinical competence,<br>writing daily medical records,<br>writing medical summaries<br><u>Outcome measures</u><br>9-point self-assessment Likert scale 1 (extremely poor) to 9 (extremely good) | <u>Study design</u><br>Descriptive study<br>Pre-test / Post test<br><u>Type of analysis</u><br>Analytical statistics<br>Mean scores<br><u>Quality appraisal rating</u><br>Score of 3 out of 7<br><u>Confidence evaluation</u><br>Competency– Very low | Students indicated improvement across all nine competency domains which were all significant at p<0.05 |
| Kim et al., 2020<sup>28</sup><br>South Korea<br>Remote teaching for medical undergraduates<br>e-Teaching and Learning System<br>Pre-recorded video lectures or live-streamed using video communication software<br>Platforms not specified | <u>Participants</u><br>Academic years 2017/2018 (n=149 to 152) sitting exams (year of study ns)<br>Academic year 2018/2019 (n=147 to 158) sitting exams (year of study ns)<br>Academic year 2019/2020 (n=143 to 145) sitting exams (year of study ns)<br><u>Outcome</u><br>Knowledge<br>Anatomy, biochemistry, histology, gastrointestinal system, respiratory system, circulatory system<br><u>Outcome measures</u><br>Examination scores | <u>Study design</u><br>Descriptive study<br>Post-test<br><u>Type of analysis</u><br>Analytical statistics<br>Mean scores<br>Comparison across three academic years<br><u>Quality appraisal rating</u><br>3 out of 7<br><u>Confidence evaluation</u><br>Knowledge– Low | <u>Knowledge (Mean±SD)</u><br>Anatomy<br>2018 (86.0±7.0); 2019 (88.1±10.3); 2020 (82.0±11.5), p<0.001<br>Effect size 2019 & 2019 compared 2020, p=-0.5150<br>Biochemistry<br>2018 (79.7±11.5); 2019 (70.9±17.3); 2020 (74.1±17.3), p<0.001<br>Effect size 2019 & 2019 compared 2020 = -0.0754<br>Histology<br>2018 (86.2±6.7); 2019; (85.1±12.9); 2020 (83.4±12.0), p=0.0754<br>Effect size 2019 & 2019 compared 2020 = -0.2127<br>Gastrointestinal system<br>2018 (86.6±8.8); 2019 (88.4±10.5); 2020 (85.9±10.4), p=-0.0825<br>Effect size 2019 & 2019 compared 2020 = -0.1605<br>Respiratory system<br>2018; (78.7±13.1); 2019 (88.2±9.2); 2020 (76.9±11.7); p<0.0001<br>Effect size 2019 & 2019 compared 2020 = -0.5504<br>Circulatory system<br>2018 (79.2±10.6); 2019 80.1±10.5; 2020 (77.3±12.1), p=0.0854<br>Effect size 2019 & 2019 compared 2020 =-0.2116 |
| Nathaniel and Black, 2021<sup>29</sup><br>USA<br>Remote, blended learning approach for teaching neuroanatomy<br>Neuroanatomical interactive virtual activities<br>“Digital Neuroanatomy” software<br>Lectures<br>Recorded on WebEx/Panopto and posted online on the Canvas platform<br>4 weeks | <u>Participants</u><br>Academic year 2019/2020 First years n=103) and 2020 (n=104)<br>Academic year 2020/2021 First years (n=104)<br><u>Outcome</u><br>Knowledge<br><u>Outcome measures</u><br>Weekly laboratory quizzes<br>Final laboratory examinations | <u>Study design</u><br>Descriptive study<br>Post-test<br><u>Type of analysis</u><br>Analytic statistics<br>Mean scores<br>Comparison across two academic years<br><u>Quality appraisal rating</u><br>5 out of 7<br><u>Confidence evaluation</u><br>Knowledge – Very low | <u>Knowledge (Mean±SD)</u><br>Final laboratory summative examination<br>2019 (92± 0.15); 2020 (90± 0.11), p=0.009 |
| Redinger and Greene, 2021<sup>33</sup><br>USA | <u>Participants</u><br>Academic year 2019/2020<br>Traditional rotation | <u>Study design</u><br>Descriptive study<br>Post test | <u>Knowledge (Mean±SD)</u> |
| Virtual clerkship in emergency medicine<br>Microsoft Teams platform for video conferences, news feed with chat functions, class assignments, daily quizzes, and grade book.<br>Simulated patient encounters employing Online MedEd Case X (Online MedEd, Austin, TX) videos and Emergency Medicine Reviews and Perspectives (EM:RAP) podcast audio of emergency medicine patients and relevant cases<br>4 weeks | Fourth years (Clerkship) (n=48)<br>Academic year 2020/2021<br>Virtual rotation<br>Fourth years (Clerkship) (n=56)<br><u>Outcome</u><br>Knowledge<br><u>Outcome measures</u><br>Emergency medicine shelf exam | <u>Type of analysis</u><br>Analytical statistics<br>Mean scores<br>Comparison across two academic years<br><u>Quality appraisal rating</u><br>4 out of 7<br><u>Confidence evaluation</u><br>Knowledge – Very low | Virtual rotation ( 81.18± 6.55);<br>Traditional rotation (79.38±6.85), p=0.174, 95% CI [-0.808, 4.415]. |
| Totlis et al., 2021<sup>30</sup><br>Greece<br>Musculoskeletal system anatomy and neuroanatomy<br>Skype for Business; the university platform Meducator. Structural specimens replaced by photographs<br>5 weeks<br>Online or pre-recorded theoretical lectures and laboratory lectures | <u>Participants</u><br>Academic year 2018/2019<br>In-Person<br>First years studying musculoskeletal anatomy (n=252)<br>Second years studying neuroanatomy (n=211)<br>Academic year 2019/2020<br>Virtual<br>First years studying musculoskeletal anatomy (n=272)<br>Second years studying neuroanatomy (n=295)<br><u>Outcomes</u><br>Knowledge<br><u>Outcome measures</u><br>Exam grades<br>Exam grades compared with previous year (2018/2019) when traditional teaching was used (face to face including practical sessions, anatomical models, cadaveric bones etc) | <u>Study design</u><br>Descriptive study<br>Post-test<br><u>Type of analysis</u><br>Analytical statistics<br>Mean scores<br>Comparison between remote and in person learning across two academic years<br><u>Quality appraisal rating</u><br>Score of 4 out of 7<br><u>Confidence evaluation</u><br>Knowledge – Very low | <u>Knowledge (Mean±SD)</u><br>Musculoskeletal anatomy:<br>In-Person ( 6.88±2.12); Virtual (6.59±1.67), p<0.001<br>Neuroanatomy<br>In-Person (6.10±2.23); Virtual (5.70±1.61), p<0.001 |
| Rosenthal et al., 2020<sup>25</sup><br>USA<br>Peer led online learning course in emergency medicine<br>Course content (8 topics) organised by 12 rising fourth-year medical students under supervision of faculty mentor/Director for Undergraduate Medical Education<br>Online Video Conferencing software | <u>Participants</u><br>Academic year 2019/2020<br>Fourth years (n=61)<br><u>Outcomes</u><br>Confidence (Comfort)<br>Imaging<br>Chest pain and EKG<br>Stroke and lumbar puncture<br>Abdominal pain<br>Altered mental status and toxicology<br>Shortness of breath and ventilators<br>Shock and sepsis<br>Trauma and FAST Exams | <u>Study design</u><br>Descriptive study<br>Pre-test / Post-test<br><u>Type of analysis</u><br>Analytic statistics<br>Mean scores<br><u>Quality appraisal rating</u><br>Score 4 out of 7<br><u>Confidence evaluation</u><br>Confidence– Very low | Mean confidence scores improved across all learning objectives (p<0.05) |
| Pre-lectures and lectures made use of:<br>Podcasts; Publications, Clinical vignettes, Online content reviews, Video conferencing<br>Platforms not specified | <u>Outcome Measures:</u><br>Self-assessments using a 5-point Likert scale of 1-5, ranging from “very uncomfortable” to “very comfortable.” |  |  |
| Suppan et al., 2021<sup>32</sup><br>Switzerland<br>Asynchronous distance learning of the National Institutes of Health Stroke Scale<br>Web-based platform e-learning module interactive content, including gamified modules and serious games, which can be accessed on regular computers as well as on smartphones and tablet compared to standard video based learning | <u>Participants</u><br>Academic year 2019/2020<br>Fifth years (75/158, rr 47.5%)<br>E learning module (n=41)<br>Video group (n=34)<br><u>Outcomes</u><br>Knowledge<br><u>Outcome measures</u><br>50-question quiz | <u>Study design</u><br>RCT<br>Intervention group<br>E-Learning module<br>Control group<br>Video<br><u>Type of analysis</u><br>Analytical statistics<br>Mean scores<br><u>Quality appraisal rating</u><br>Score of 7 out of 11<br><u>Confidence evaluation</u><br>Moderate | Overall quiz score (Mean±SD)<br>e-learning module (38±3, 95% CI 39); video group (35±3, 95% CI 34-36), p<0.001 |
| Schmitz et al., 2021<sup>31</sup><br>Germany<br>Surgical online learning platform<br>Interactive online platform to teach operative techniques and skills. Surgical procedures were videorecorded in our operating theatre and processed in order to design an interactive video format<br>Seven educational sessions | <u>Participants</u><br>Academic year ns<br>(n=44/58 completed the study)<br>Second years (82%)<br>Intervention group (n=21)<br>Control group (n=23)<br><u>Outcomes</u><br>Knowledge<br><u>Outcome measures</u><br>Online exam consisting of 10 multiple choice questions | <u>Study design</u><br>RCT<br>Intervention group<br>Video based preparation<br>Control group<br>Textbook based preparation<br><u>Type of analysis</u><br>Analytical statistics<br>Percentage of correct, incorrect and ‘don’t know’ choices<br><u>Quality appraisal rating</u><br>Score of 11 out of 11<br><u>Confidence evaluation</u><br>Very Low | Percentage of correct choices<br>Intervention group:(0.67±0.02);<br>Control group (0.60±0.02), p=0.001<br>Percentage of incorrect choices<br>Intervention group (0.24±0.19);<br>Control group (0.29 ± 0.223); p=0.04 |
| Pang et al., 2021<sup>23</sup><br>USA<br>An Informed Consent activity module within a virtual surgical clerkship<br>A pre-recorded lecture with presentation slides<br>A videoconference with 3 students, 2 standardised patients and a facilitator to practice obtaining informed consent for a common surgical procedure | <u>Participants</u><br>Academic year 2019/2020<br>Third years (34/ 90; 38%) who completed the module and took part in the evaluation<br><u>Outcomes</u><br>Competency in 4 domains:<br>The ability to identify the key elements of informed consent<br>The ability to describe common challenges in the informed consent process | <u>Study design</u><br>Descriptive study<br>Pre-test / Post-test (retrospective)<br><u>Type of analysis</u><br>Analytical statistics<br>Mean scores<br><u>Quality appraisal rating</u><br>Score 3 out of 7<br><u>Confidence evaluation</u><br>Competency – Very low | Results for 4 domains: (Mean±SD)<br>Identifying the elements of informed consent:<br>Pre-test (1.9±1.4);<br>Post-test (3.5±0.93), p<0.001<br>Describing common challenges in informed consent:<br>Pre-test (1.0±1.15);<br>Post-test (3.3±0.90 ), p<0.001<br>Applying NM-CCS quality framework:<br>Pre-test (2.1±1.24);<br>Post-test (3.5±0.66), p<0.001<br>Documenting informed consent: |
| Platforms not specified | The ability to apply the recommended quality framework (NM-CCS)<br>The ability document informed consent.<br><u>Outcome measure</u><br>Self-assessment 6-point scale (0 being none/no competence and 5 being an extremely high level of competence) |  | Pre-test (2.0±1.19);<br>Post-test (3.4±0.61), p<0.001 |

Key: EKG : Electrocardiogram; FAST: Focused Assessment with Sonography for Trauma; NM-CCS: New Mexico Clinical Communication Scale; RCT: Randomised Controlled Trial
<sup>a</sup> High-fidelity simulation refers to simulation experiences that are extremely realistic and provide a high level of interactivity and realism for the learner

These covered a wide range of both university and clinical based modules/ courses and included neurosurgery,^21^ surgical instruments, knot tying and suturing,^24^ digital histology,^26^ a residency preparation course,^22^ simulated patient consultations, documentation, and case presentation,^27^ simulated clinical experience in respiratory unit and general medicine,^20^ generic medical education,^28^ neuroanatomy,^29^ emergency medicine^25,33^ musculoskeletal system anatomy and neuroanatomy,^30^ the National Institutes of Health Stroke Scale,^32^ operative techniques and skills,^31^ and informed consent for surgical procedures.^23^

A variety of different online platforms was used to deliver synchronous learning; five used the Zoom video conferencing platform^20,21,24,26,27^ three used the University Supported Management Systems: CANVAS^22,29^ or Meducator,^30^ one used Microsoft teams,^33^ another Skype for business,^30^ and three did not specify the type of video communication software used.^23,25,28^ Other methods included neuroanatomical interactive virtual activities using “Digital Neuroanatomy” software,^29^ simulated patient encounters employing online MedEd Case X videos,^33^ and structural specimens replaced by photographs.^30^ Five studies also incorporated asynchronous elements using pre-recorded lectures^23,28,30^ or readily available podcasts.^25,33^ For one further study the course content (8 topics) was organised by 12 rising^1^ fourth-year medical students under supervision.^25^ The two RCTs used bespoke interactive online platforms^31,32^ and compared the outcomes to those students learning the same topic via a standard video format^31^ or textbook based preparation.^32^

Studies were conducted with students in their final year (Clerkship / Interns) (n=7),^20,22,24,25,27,32,33^ first year (n=2),^29,30^ second and third years (n=1),^26^ third year only (n=1),^23^ across all years (n=1),^21^ and a further two did not specify the year of study.^28,31^ Outcomes explored were confidence (n= 5),^21,22,24,25,27^ competency (n=2)^20,23^ and knowledge (n=6).^26,28–30,32,33^

Levels of competency, confidence and knot tying and suturing skills were found to have improved across the course of learning and a further study suggested that levels of competency were the same when learning was conducted virtually (2020) compared to in-person pre-COVID (2019). Evidence from RCTs showed that knowledge was greater when learning was conducted using bespoke interactive platforms with a standard video format reported during the COVID pandemic. Evidence from descriptive studies showed mixed results for knowledge, assessed and compared between cohorts at the end of virtual learning (2020) and in-person learning (2019). Four studies reported lower levels of knowledge for students in the virtual cohort and one further study found no difference.

### Overview of the evidence base for dental students

Two post-test descriptive studies (see Table 5) conducted in Poland^34^ and Germany^35^ provided evidence of the effectiveness of alternative education delivery strategies for undergraduate dental students studying specific modules or courses in conservative dentistry with endodontics^34^ or operative dentistry^35^ during the COVID-19 pandemic. In one study, the teaching consisted of asynchronous online screencasts (screen-captured PowerPoint presentations with narrated audio) using Stud-IP (a source learning management system) and discussed via synchronous video meetings using the Zoom video videoconferencing platform.^35^ The other study used a blended learning approach using the Blackboard Collaborate platform.^34^ The outcome of interest across both studies was knowledge acquisition. Findings from both studies suggest that these alternative educational methods contributed towards knowledge and skill acquisition assessed through a self-assessment survey completed by fourth year students^34^ and through a final summative examination.^35^ However, the evidence suggests lower levels of knowledge for the subtopic of periodontology and lower levels of practical skills for 3^rd^ year dental students when learning was conducted virtually compared to in-person.

**Table 5:** Characteristics of included studies focusing on dental students.

| Author/s<br>Country | Participants | Study design<br>Type of analysis | Findings |
| --- | --- | --- | --- |
| Focus<br>Remote platform | Outcomes / Outcome measures | Quality appraisal rating |  |
| Nijakowski et al., 2021<sup>34</sup><br>Poland<br>Blended learning in conservative dentistry with endodontics<br>Blackboard Collaborate<br>2019/2020<br>Online classes<br>2021/2021<br>Full blended learning, clinical classes, e-learning seminars, and online meetings via Microsoft teams | <u>Participants</u><br>Academic year 2019/2020<br>Third years<br>Clinical classes (n=39)<br>Online only classes (n=35)<br>Academic years 2020/2021<br>Fourth years (n=74)<br><u>Outcomes</u><br>Theoretical knowledge, practical skills, and interpersonal skills<br><u>Outcome measures</u><br>5-point self-assessment Likert scales | <u>Study design</u><br>Descriptive study<br>Post test<br><u>Type of analysis</u><br>Analytic statistics<br>Mean scores<br>Comparison between remote and in person learning within the same academic year<br>Comparison between academic years (retrospective self-assessment during the third year compared to fourth year)<br><u>Quality appraisal rating</u><br>Score 4 out of 7<br><u>Confidence evaluation</u><br>Knowledge – Very low<br>Skills – Very low | <u>Theoretical knowledge</u> (Mean: Q<sub>1</sub>-Q<sub>3</sub>)<br>3<sup>rd</sup> year (retrospective) 3.0 (3.0 -4.0); 4<sup>th</sup> Year 4.0 (4.0-4.0), p=0.001<br>3<sup>rd</sup> year (retrospective) In-Person 3.0 (3.0-4.0); 3<sup>rd</sup> year (retrospective) Virtual 3.0 (3.0-4.0), p=0.702<br>4<sup>th</sup> year In-Person 4.0 (4.0-4.0); 4<sup>th</sup> year Virtual 4.0 (4.0-4.0), p=0.879<br><u>Practical skills</u><br>3<sup>rd</sup> year (retrospective) 3.0 (2.0-4.0); 4<sup>th</sup> Year 4.0 (3.0-4.0), p<0.001<br>3<sup>rd</sup> year (retrospective) In-Person 3.0 (2.0-4.0); 3<sup>rd</sup> year (retrospective) Virtual 2.0 (1.0-2.0), p<0.001<br>4<sup>th</sup> year In-Person Year 4.0 (3.0-4.0), 4<sup>th</sup> year Virtual 3.0 (3.0-4.0), p=0.083<br><u>Interpersonal skills</u><br>3<sup>rd</sup> year (retrospective) 4.0 (3.0-5.0); 4<sup>th</sup> Year 4.0 (4.0-5.0), p=0.048<br>3<sup>rd</sup> year (retrospective) In-Person 4.0 (3.0-5.0); 3<sup>rd</sup> year (retrospective) Virtual 3.0 (2.0-4.0), p=0.008<br>4<sup>th</sup> year In-Person 4.0 (4.0-5.0), 4<sup>th</sup> year Virtual 4.0 (4.0-5.0), p=0.952 |
| Kanzow et al., 2021<sup>35</sup><br>Germany<br>Preclinical phantom course in operative dentistry<br>Theoretical knowledge was taught via screen-captured PowerPoint presentations with narrated audio)<br>Stud.IP, an open-source learning management system by using a MediaCast plugin<br>3 a week for 10 weeks<br>Live and interactive video meetings using Zoom video conferencing platform<br>Physical skills taught onsite using phantom heads with natural tooth model | <u>Participants</u><br>Summer term 2020<br>Students enrolled in the pre-clinical phantom course in operative dentistry (n=33)<br>31 students were eligible to take the final exam<br><u>Outcomes</u><br>Knowledge<br>Cariology, restorative dentistry and, preventative dentistry, endodontology and periodontology<br><u>Outcome measures</u><br>Summative electronic examination of theoretical knowledge. 30 equally-weighted questions including multiple choice, true/false and open-ended items. A fixed pass mark of 60%. Students had to perform a pre-defined number of treatments in the physical skills part of the course to be admitted to the exam | <u>Study design</u><br>Descriptive study<br>Post-test<br><u>Analytical statistics</u><br>Mean scores<br>Comparison of scores between topics<br><u>Quality appraisal rating</u><br>Score 4 out of 7<br><u>Confidence evaluation</u><br>Knowledge - Low | <u>Knowledge</u><br>Credit (%) awarded in each topic (mean±SD)<br>Cariology, Restorative Dentistry and Preventive Dentistry: 75.8+34.5<br>Endodontology: 79.2+31.2<br>Periodontology: 58.9+37.2<br>Overall credit: 74.5+34.6<br>Examination items in periodontology showed inferior results compared with other topics (p<.001) |

Key: Q: quartiles

### Overview of evidence base for nursing students

Three descriptive studies (see Table 6) conducted in Spain,^36^ Japan^37^ and USA^38^ provided evidence for the effectiveness of alternative educational delivery strategies for nursing students studying a specific module in human genomics,^37^ simulation in paediatric clinical practice^38^ and for the delivery of remote OSCEs with COPD patients^36^ during the COVID-19 pandemic. All three studies compared a group of students receiving a remotely delivered educational package with a group receiving standard, in-person education. In two studies the comparison groups were students from the previous, pre-COVID academic year, however, Weston and Zauche^38^ studied a cohort of students from the same academic year, 2019-2020, where half had received the standard educational package before the alternative version was introduced. Only one study used a pre-test / post-test design and thus compared results within as well as between groups.^37^ In this study, the conventional course was transferred to remote synchronous learning (narrative over PowerPoint), also uploading handouts and worksheets with no changes to content.^37^ Arrogante et al.^36^ used the virtual classroom platform Blackboard Collaborate to conduct OSCEs comprising eight simulated clinical scenarios with standardised patients. Weston and Zauche^38^ substituted virtual simulation using the i-Human platform to replace in-person clinical practice and simulation laboratory learning. Outcomes explored were competency (n=2)^36,37^ confidence (n=1),^37^ and knowledge (n=2).^37,38^

**Table 6:**
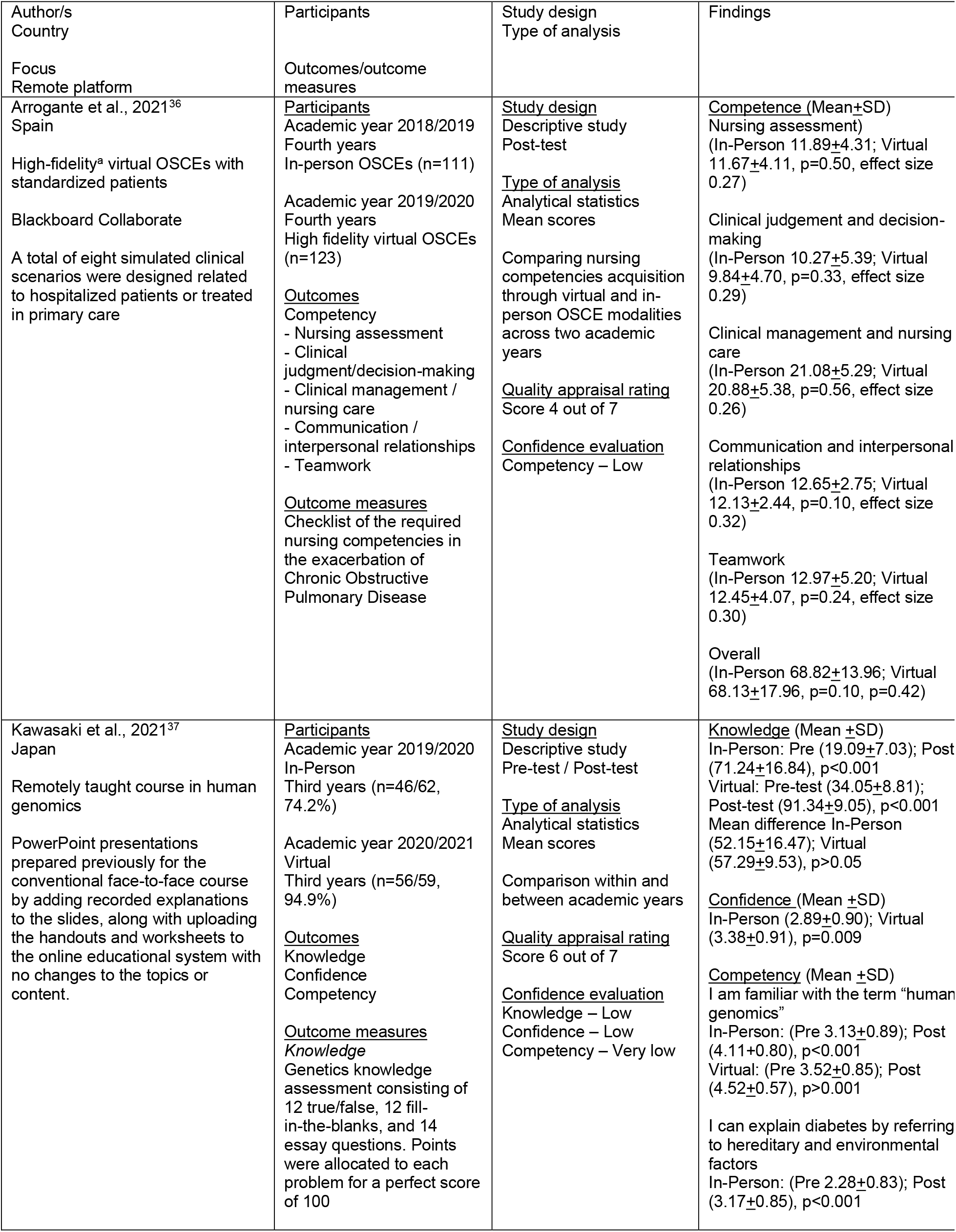
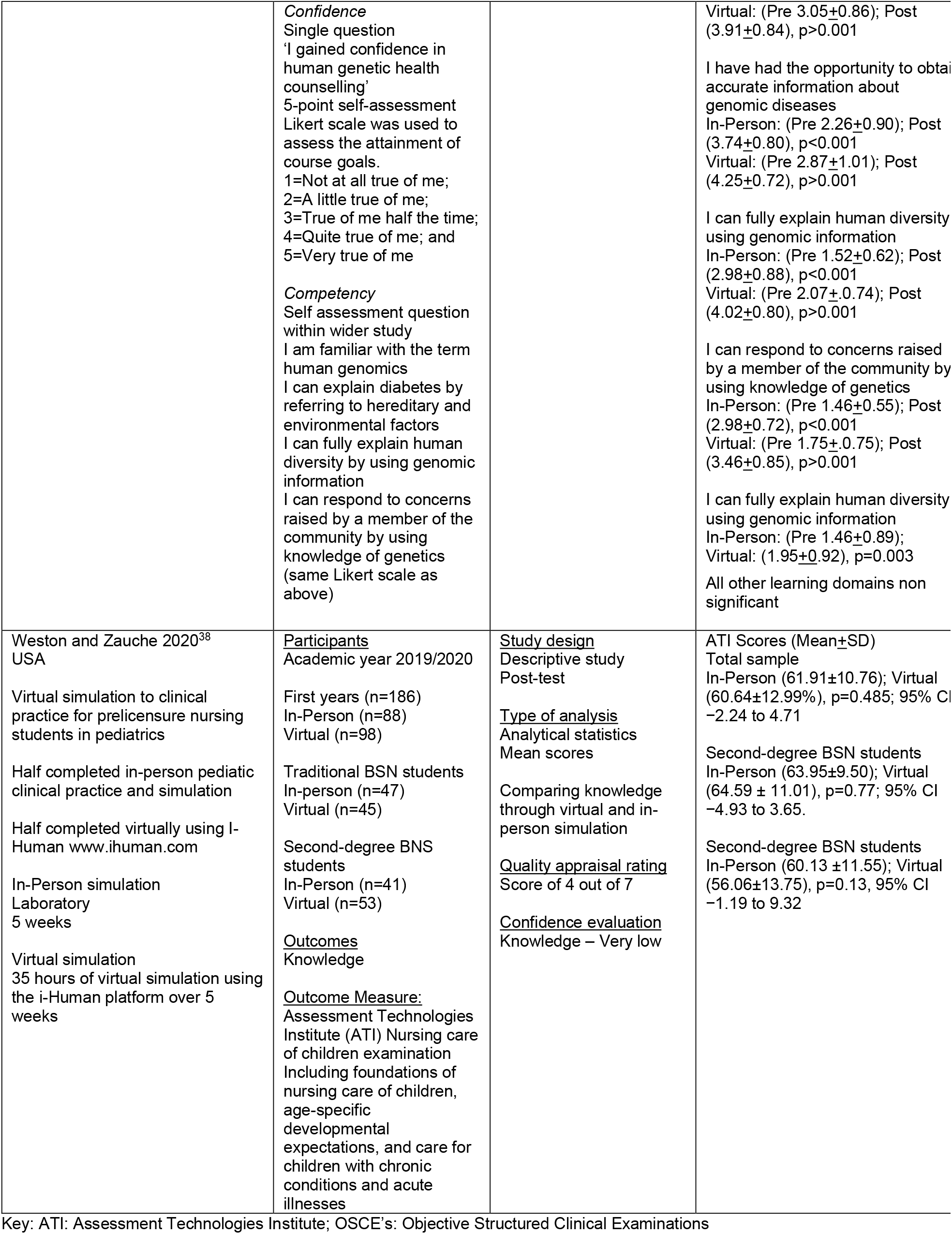
Characteristics of included studies focusing on nursing students.

| Author/s<br>Country | Participants | Study design<br>Type of analysis | Findings |
| --- | --- | --- | --- |
| Focus<br>Remote platform | Outcomes/outcome<br>measures |  |  |
| Arrogante et al., 2021<sup>36</sup><br>Spain<br>High-fidelity<sup>a</sup> virtual OSCEs with standardized patients<br>Blackboard Collaborate<br>A total of eight simulated clinical scenarios were designed related to hospitalized patients or treated in primary care | <u>Participants</u><br>Academic year 2018/2019<br>Fourth years<br>In-person OSCEs (n=111)<br>Academic year 2019/2020<br>Fourth years<br>High fidelity virtual OSCEs (n=123)<br><u>Outcomes</u><br>Competency<br>- Nursing assessment<br>- Clinical judgment/decision-making<br>- Clinical management / nursing care<br>- Communication / interpersonal relationships<br>- Teamwork<br><u>Outcome measures</u><br>Checklist of the required nursing competencies in the exacerbation of Chronic Obstructive Pulmonary Disease | <u>Study design</u><br>Descriptive study<br>Post-test<br><u>Type of analysis</u><br>Analytical statistics<br>Mean scores<br>Comparing nursing competencies acquisition through virtual and in-person OSCE modalities across two academic years<br><u>Quality appraisal rating</u><br>Score 4 out of 7<br><u>Confidence evaluation</u><br>Competency – Low | <u>Competence (Mean±SD)</u><br>Nursing assessment)<br>(In-Person 11.89±4.31; Virtual 11.67±4.11, p=0.50, effect size 0.27)<br>Clinical judgement and decision-making<br>(In-Person 10.27±5.39; Virtual 9.84±4.70, p=0.33, effect size 0.29)<br>Clinical management and nursing care<br>(In-Person 21.08±5.29; Virtual 20.88±5.38, p=0.56, effect size 0.26)<br>Communication and interpersonal relationships<br>(In-Person 12.65±2.75; Virtual 12.13±2.44, p=0.10, effect size 0.32)<br>Teamwork<br>(In-Person 12.97±5.20; Virtual 12.45±4.07, p=0.24, effect size 0.30)<br>Overall<br>(In-Person 68.82±13.96; Virtual 68.13±17.96, p=0.10, p=0.42) |
| Kawasaki et al., 2021<sup>37</sup><br>Japan<br>Remotely taught course in human genomics<br>PowerPoint presentations prepared previously for the conventional face-to-face course by adding recorded explanations to the slides, along with uploading the handouts and worksheets to the online educational system with no changes to the topics or content. | <u>Participants</u><br>Academic year 2019/2020<br>In-Person<br>Third years (n=46/62, 74.2%)<br>Academic year 2020/2021<br>Virtual<br>Third years (n=56/59, 94.9%)<br><u>Outcomes</u><br>Knowledge<br>Confidence<br>Competency<br><u>Outcome measures</u><br><u>Knowledge</u><br>Genetics knowledge assessment consisting of 12 true/false, 12 fill-in-the-blanks, and 14 essay questions. Points were allocated to each problem for a perfect score of 100 | <u>Study design</u><br>Descriptive study<br>Pre-test / Post-test<br><u>Type of analysis</u><br>Analytical statistics<br>Mean scores<br>Comparison within and between academic years<br><u>Quality appraisal rating</u><br>Score 6 out of 7<br><u>Confidence evaluation</u><br>Knowledge – Low<br>Confidence – Low<br>Competency – Very low | <u>Knowledge (Mean ±SD)</u><br>In-Person: Pre (19.09±7.03); Post (71.24±16.84), p<0.001<br>Virtual: Pre-test (34.05±8.81); Post-test (91.34±9.05), p<0.001<br>Mean difference In-Person (52.15±16.47); Virtual (57.29±9.53), p>0.05<br><u>Confidence (Mean ±SD)</u><br>In-Person (2.89±0.90); Virtual (3.38±0.91), p=0.009<br><u>Competency (Mean ±SD)</u><br>I am familiar with the term “human genomics”<br>In-Person: (Pre 3.13±0.89); Post (4.11±0.80), p<0.001<br>Virtual: (Pre 3.52±0.85); Post (4.52±0.57), p>0.001<br>I can explain diabetes by referring to hereditary and environmental factors<br>In-Person: (Pre 2.28±0.83); Post (3.17±0.85), p<0.001 |
|  | *Confidence*<br>Single question<br>'I gained confidence in human genetic health counselling'<br>5-point self-assessment Likert scale was used to assess the attainment of course goals.<br>1=Not at all true of me;<br>2=A little true of me;<br>3=True of me half the time;<br>4=Quite true of me; and<br>5=Very true of me<br>*Competency*<br>Self assessment question within wider study<br>I am familiar with the term human genomics<br>I can explain diabetes by referring to hereditary and environmental factors<br>I can fully explain human diversity by using genomic information<br>I can respond to concerns raised by a member of the community by using knowledge of genetics (same Likert scale as above) |  | Virtual: (Pre 3.05±0.86); Post (3.91±0.84), p>0.001<br>I have had the opportunity to obtain accurate information about genomic diseases<br>In-Person: (Pre 2.26±0.90); Post (3.74±0.80), p<0.001<br>Virtual: (Pre 2.87±1.01); Post (4.25±0.72), p>0.001<br>I can fully explain human diversity using genomic information<br>In-Person: (Pre 1.52±0.62); Post (2.98±0.88), p<0.001<br>Virtual: (Pre 2.07±0.74); Post (4.02±0.80), p>0.001<br>I can respond to concerns raised by a member of the community by using knowledge of genetics<br>In-Person: (Pre 1.46±0.55); Post (2.98±0.72), p<0.001<br>Virtual: (Pre 1.75±0.75); Post (3.46±0.85), p>0.001<br>I can fully explain human diversity using genomic information<br>In-Person: (Pre 1.46±0.89); Virtual: (1.95±0.92), p=0.003<br>All other learning domains non significant |
| Weston and Zauche 2020<sup>38</sup><br>USA<br>Virtual simulation to clinical practice for prelicensure nursing students in pediatrics<br>Half completed in-person pediatric clinical practice and simulation<br>Half completed virtually using I-Human www.ihuman.com<br>In-Person simulation<br>Laboratory<br>5 weeks<br>Virtual simulation<br>35 hours of virtual simulation using the i-Human platform over 5 weeks | <u>Participants</u><br>Academic year 2019/2020<br>First years (n=186)<br>In-Person (n=88)<br>Virtual (n=98)<br>Traditional BSN students<br>In-person (n=47)<br>Virtual (n=45)<br>Second-degree BSN students<br>In-Person (n=41)<br>Virtual (n=53)<br><u>Outcomes</u><br>Knowledge<br><u>Outcome Measure:</u><br>Assessment Technologies Institute (ATI) Nursing care of children examination<br>Including foundations of nursing care of children, age-specific developmental expectations, and care for children with chronic conditions and acute illnesses | <u>Study design</u><br>Descriptive study<br>Post-test<br><u>Type of analysis</u><br>Analytical statistics<br>Mean scores<br>Comparing knowledge through virtual and in-person simulation<br><u>Quality appraisal rating</u><br>Score of 4 out of 7<br><u>Confidence evaluation</u><br>Knowledge – Very low | ATI Scores (Mean±SD)<br>Total sample<br>In-Person (61.91±10.76); Virtual (60.64±12.99%), p=0.485; 95% CI -2.24 to 4.71<br>Second-degree BSN students<br>In-Person (63.95±9.50); Virtual (64.59 ± 11.01), p=0.77; 95% CI -4.93 to 3.65.<br>Second-degree BSN students<br>In-Person (60.13 ±11.55); Virtual (56.06±13.75), p=0.13, 95% CI -1.19 to 9.32 |

Key: ATI: Assessment Technologies Institute; OSCE's: Objective Structured Clinical Examinations

The evidence suggests that levels of competency were the same and levels of confidence were higher when learning or assessment was conducted virtually (2020) compared to in-person pre-COVID (2019). Knowledge improves regardless of whether the learning has been conducted virtually (2020) or in-person pre-COVID (2019).

### Overview of the evidence base for pharmacy students

Four descriptive studies (see Table 7), all conducted in the USA, provided evidence for the effectiveness of alternative education delivery strategies for undergraduate pharmacy students studying specific modules or courses in integrated patient care,^39^ hypertension/drug information,^40^ advanced pharmacy experience,^41^ delivery of remote Objective Structured Clinical Examinations (OSCEs) for patient counselling, and taking a medical history^42^ during the COVID-19 pandemic. Two studies used a pre-test/post-test design,^40,41^ the remaining two reported a post-test only study design, with a comparison between the study population and an earlier (pre-COVID) cohort of students.^39,42^

**Table 7:** Characteristics of included studies focusing on pharmacy students.

| Author/s<br>Country | Participants | Study design<br>Type of analysis | Findings |
| --- | --- | --- | --- |
| Focus<br>Remote platform | Outcomes / Outcome measures |  |  |
| Phillips et al., 2021<sup>39</sup><br>USA<br>Remote delivery of Integrated Patient Care Capstone course<br>Zoom video conferencing platform<br>60% of the course competed in-person before transitioning to remote learning which consisted of weekly class sessions | <u>Participants</u><br>Academic year 2019/2020<br>In-person<br>Third (n=134)<br>Academic year 2020/2021<br>60% course completed in person before moving to remote learning<br>Third years (n=126)<br><u>Outcomes</u><br>Drug therapy knowledge<br>Application of drug therapy guidelines<br>Improving clinical reasoning, strengthening pharmacists' patient care process, skill development<br><u>Outcome measures</u><br>*Knowledge / performance:*<br>Quizzes<br>Mid-term examination result<br>Final examination results<br>*Competency & confidence:*<br>6-item self-assessment scale | <u>Study design</u><br>Descriptive study<br>Post test<br><u>Type of analysis</u><br>Analytical statistics<br>Mean scores<br>Comparison between remote and in person learning within the same academic year<br>Comparison between two academic years<br><u>Quality appraisal rating</u><br>Score 3 out of 7<br><u>Confidence evaluation</u><br>Knowledge – Very Low<br>Confidence - Low<br>Competency – Low | <u>Knowledge</u><br>Quiz average (Mean ±SD)<br>2019 cohort (23.0±3.0);<br>2020 cohort (23.6±1.9), $p>0.05$)<br>2020 Spring semester<br>In-Person (7.7 ± 1.8);<br>2020 summer semester<br>Virtual (8.2 ± 1.6), $p<0.05$)<br>Mid-term examination (Mean ±SD)<br>2019 cohort (21.3±4.8);<br>2020 cohort 22.1±5.0, $p>0.05$)<br>Final examination (Mean ±SD)<br>2019 cohort (23.1±5.4);<br>2020 cohort 21.3±5.4, $p<0.01$)<br>2020 Spring semester<br>In-Person (23.1 ± 5.4),<br>2020 summer semester<br>Virtual (21.3 ± 5.4); $p<0.05$)<br><u>Competency</u><br>No significant difference in self-assessed skill development when compared between 2019 and 2020 using anonymous course evaluation data (Mann-Whitney U test; $p>0.05$)<br><u>Confidence</u><br>No significant associations were found between level of student confidence in skill development and performance on final practical exam or in the overall course in 2020 (Spearman Correlation test, $p>0.05$) |
| Cowart and Updike 2021<sup>40</sup><br>USA<br>Remote delivery of a hypertension/drug information simulation-based learning<br>Blackboard Collaborate<br>Across 3 days after 1.5 hours didactic lectures and 2.5 hours laboratory instructive session, pre case vignettes | <u>Participants</u><br>Academic year 2019/2020<br>First years (n=87)<br>Response rate pre-test (95%)<br>Response rate post test (62%)<br><u>Outcomes</u><br>Blood pressure techniques<br>Application of drug information<br>Assessment of communication skills<br><u>Outcome measures</u><br>*Competency*<br>4-point self-assessment Likert scale<br>(1=strongly disagree, 2=disagree, 3=agree, 4=strongly agree)<br>*Confidence*<br>5-point self-assessment Likert-scale | <u>Study design</u><br>Descriptive study<br>Pre-test / Post-test<br><u>Type of analysis</u><br>Analytical statistics<br>Mean scores<br><u>Quality appraisal rating</u><br>Score 3 out of 7<br><u>Confidence evaluation</u><br>Confidence - Low<br>Competency – Very low | <u>Confidence</u> (Mean ±SD)<br>Blood pressure techniques<br>(Pre 2.75±0.99; Post 4.13±0.7, $p<0.001$)<br>Application of drug information<br>(Pre 3.55±1.06; Post 4.39±0.81; $p=0.002$)<br>Assessment of communication skills<br>(Pre 3.05±0.99; Post 3.87±0.83) $p<0.001$)<br><u>Competency</u> (Mean ±SD)<br>Blood pressure techniques<br>(Pre 3.28±0.57, Post 3.22±0.67, $p=0.859$)<br>Application of drug information<br>(Pre 3.17±0.51, Post 3.30±0.66, $p=0.864$)<br>Assessment of communication |
|  | (0=not at all confident, 1=slightly confident, 2=somewhat confident, 3=moderately confident, 4=very confident) |  | (Pre 3.17±0.51, post 3.44±0.54, p=0.007) |
| Scoular et al., 2021<sup>42</sup><br>USA<br>Remote delivery of OSCEs in patient counselling and taking a medical history<br>Zoom video conferencing platform | <u>Participants</u><br>Academic year 2019/2020<br>First years (n=144)<br>Academic years 2020/2021<br>First years (n=106)<br><u>Outcomes</u><br>Skills (Patient centred communication; empathy; trust; professionalism; general verbal and non-verbal communication skills)<br><u>Outcome measures</u><br>Cumulative OSCE<br>Patient centred communication OSCE<br>Students were required to counsel a standardized patient on two prescription products with unique dosage forms (e.g., inhalers). Students' skills were graded by standardized patients | <u>Study design</u><br>Descriptive study<br>Post test<br><u>Type of analysis</u><br>Analytical statistics<br>Mean scores<br>Comparison between remote and in person learning<br>Comparison of performance scores between two academic years<br><u>Quality appraisal rating</u><br>Score 5 out of 7<br><u>Confidence evaluation</u><br>Knowledge – Very low | <u>Patient centred communication OSCE</u><br>Overall score (Median, range)<br>2019 (96.47, 36.47);<br>2020 (99.00, 23.00), p=0.000<br>effect size -0.29<br>Comparison between 2019/2020 sub domains<br>Establishing a trusting relationship (p=0.000), effect size -0.32<br>Effective verbal and non-verbal communication (p=0.001, effect size -0.21)<br>Provided patient friendly education (p=0.026, effect size -0.14)<br>Organizing the encounter (p=0.000, effect size -0.13)<br><u>Cumulative OSCE</u><br>Total variable score (Median)<br>2019 (16.00, 10.00);<br>2020 (16.00, 16.00), p=0.039, effect size -0.13<br>Comparison between 2019/2020 sub domains<br>Demonstrates empathy (p=0.245)<br>Appropriate non-verbal communication (p=0.259)<br>Professionalism (p=0.750)<br>Global feedback: Establishing Trust (p=0.015, effect size -0.15) |
| Singh et al., 2021<sup>52</sup><br>USA<br>Virtual case-based learning elective rotation for Advanced Pharmacy Experience<br>Asynchronous independent work and synchronous video conferencing<br>University Supported Management System: CANVAS<br>Zoom video conferencing platform<br>6-weeks | <u>Participants</u><br>Students (n=68/70)<br>No further details provided<br><u>Outcomes</u><br>Confidence (based on SLOs below)<br><u>Knowledge</u><br>Student Learning Outcomes (SLOs) (n=8)<br>SLO 1: Retrieve evidence-based medicine in the patient decision-making process<br>SLO 2: Evaluate and apply evidence-based medicine in the patient decision-making process<br>SLO 3: Analyse patient-specific background (i.e., informational, functional, socioeconomic, cultural, and behavioural) to establish patient-specific goals<br>SLO 4: Prepare and communicate patient care plans<br>SLO 5: Design, and redesign as appropriate, a safe, and effective patient specific plan | <u>Study design</u><br>Descriptive study<br>*Confidence*<br>Pre-test / Post test<br>*Knowledge*<br>Post-test<br><u>Type of analysis</u><br>Descriptive statistics<br>Mean scores<br><u>Quality appraisal rating</u><br>Score 4 out of 7<br><u>Confidence evaluation</u><br>Knowledge – Very Low<br>Confidence – Low | <u>Knowledge</u><br>(SLO's: mean scores)<br>SLO 1: 76.31%<br>SLO 2 80.42%<br>SLO 3 76.31%<br>SLO 4 81.14%<br>SLO 7 :75.51%<br>SLO 8: 75.77%.<br>The average score for the one graded activity mapped to SLO 5 was 76.31%<br>SLO 6 was 76.31%<br><u>Confidence</u><br>The mean difference in the student responses showed a greater than average 10-point improvement in their ability to demonstrate learning outcomes |
|  | SLO 6: Develop patient-specific monitoring plans to assess efficacy and safety<br>SLO 7: Develop drug-related education materials<br>SLO 8: Clearly communicate educational materials to preceptors and peers<br><u>Outcome Measures:</u><br>*Confidence*<br>100-point levelled ability scale with each of five levels of ability spanning a range of 0 to 20<br>*Knowledge*<br>Seven graded activities (case-based quizzes, drug consultations and presentations, journal club activities, and the closeout exams) were used to assess the achievement of SLOs, with a target minimum average of 80% as an acceptable level for achieving outcomes |  |  |

Key: OSCE's : Objective Structured Clinical Examinations; SLO: Student Learning Outcomes

In one study the teaching included remote synchronous learning,^41^ three studies used the Zoom videoconferencing platform,^39,41,42^ two studies used the University platform Blackboard Collaborate^40^ and one study also used the University Supported Management System: CANVAS.^41^ The outcomes of interest that were explored were competency (n=2),^39,40^ confidence (n=2),^40,41^ knowledge (n=2),^39,41^ skills (n=2)^41,42^

Evidence suggests competency outcomes improved across the course of learning and were similar when learning was conducted virtually (2020) compared to in-person pre-COVID (2019). Confidence was found to either improve across the course of learning or be the same for virtual (2020) compared to in-person pre-COVID (2019) learning. However, lower levels of knowledge were reported when learning was conducted virtually compared to in-person pre-COVID. The evidence suggests that, overall, students performed similarly between in-person (2019) and online (2020) OSCEs, although for some, skills performance was higher when students undertook these virtually.

## Discussion

The findings of this rapid review are based on very limited evidence for dental (2 descriptive studies), pharmacy (4 descriptive studies) and nursing (3 descriptive studies) education. Only one finding from across all twelve of the descriptive studies that focused on medical education was rated as being of moderate quality. As expected, levels of knowledge, competency and confidence improved over the course of virtual learning. However, when results were compared to students who had completed in-person learning in the years before the COVID-19 pandemic, results were mixed. Most studies across the disciplines reported similar findings across all outcome variables suggesting that virtual learning produced similar results to in-person learning. To our knowledge this is the first rapid review of the effectiveness of alternative education delivery strategies for undergraduate and postgraduate medical, dental, nursing and pharmacy education during the COVID-19 pandemic.

Previous systematic reviews showed online learning outcomes to be comparable to in-person learning. At the time of conducting this rapid review we were unable to locate any reviews that took an interdisciplinary approach. Given the potential overlap and value in sharing practices across the various healthcare educational contexts, we aimed to address this gap.

Evidence from two RCTs showed that knowledge was greater when learning was conducted using bespoke interactive platforms compared with non-interactive formats, reported during the COVID pandemic.^31,32^ These findings concur with research conducted in the field prior to COVID-19, with three systematic reviews suggesting that pre-planned online eLearning for undergraduates in health professions is equivalent, possibly superior to traditional learning.^43–45^

Data from this rapid review indicated that the transition from traditional teaching into remote methods seemed to affect students’ performance at exams, particularly so for the practical based subjects in dentistry and medicine. It is recognised that emergency remote teaching and learning differs from planned online learning.^46,47^ Most remote teaching and learning that initially took place during the COVID-19 pandemic was not planned and was adapted promptly due to the emergency circumstances that presented. In addition, this new learning did not take into account the additional stress that e-learning can cause^48^ or incorporate strategies to increase social presence which Natajaran and Joseph^49^ argue is essential to improve student nurses’ satisfaction with online teaching.

### Implications for policy and practice

Healthcare educators need to revisit the research base surrounding remote learning and consider this evidence when planning future online education. Whilst lessons learnt were quickly put into place, the COVID-19 pandemic brought issues to the fore that have long been debated in healthcare education: reduced clinical exposure, a move away from mass didactic education, and the need to ensure all healthcare students are provided with the skills and knowledge required to transition to competent caring health professionals with the ability to think critically and source and apply evidence to practice. With the increasing need for skilled healthcare professionals, policy makers need to consider how educational institutions can be provided with the resources required and how existing educators can be upskilled and supported to develop technology-enhanced learning experiences. Students from school entry age onwards need to be prepared for more online and blended learning experience which should include providing them with strategies they can use to support their emotional and psychological well-being, whilst accessing remote learning. Future research should investigate the effectiveness of blended learning approaches compared to more traditional education, in addition to investigating the views and perceptions of both students and educators and the barriers and facilitators to engaging effectively in blended learning.

## Limitations

To complete the review rapidly a limited number of databases were searched, and further studies may have been identified if additional bibliographic databases had been used. Out of the 23 included studies none was conducted within the UK and the majority (n=21) were descriptive studies. All included studies focused on undergraduate not postgraduate education. Of these, 11 studies employed a pre-test/post-test design, and the remainder were post-test only evaluations. The two RCTs both used a test or examination to assess knowledge, but these evaluated two different interventions and therefore statistical pooling of data using meta-analysis was not appropriate. Furthermore, both studies had small sample sizes and poor response rates (75/158 and 44/58).

Regarding the limitations of this review’s methods, the tool used for evaluating the confidence of the quantitative descriptive studies is an adaptation of GRADE and has not been approved by the tool’s originators. Finding well conducted comparative research proved challenging as not all educational researchers sign up to this experimental ideology when it comes to investigating teaching. Indeed, most published educational studies are small scale and qualitative in nature. There is, however, an agreement that there is a lack of high-quality studies to serve as models for future development in remote learning and teaching.^50,51^ We therefore suggest that studies that do apply the experimental approach should aim to enhance their research rigour in order for them to provide findings that can be synthesised more meaningfully. We also recognise the potential impact of the pandemic on resources and time, all of which would have likely impacted the quality of research. For this reason, we suggest that our rapid review provides a platform for further research that will consider the large body of literature that has emerged from the various fields of healthcare education since we conducted our review.

## Conclusions

Remote teaching was valued, and learning was achieved, but the comparative effectiveness of virtual versus in-person teaching delivered in a pandemic is less clear. In addition, the available evidence is insufficient to demonstrate equivalence for student speciality groups and it is unclear whether planned remote teaching, rather than relying on emergency adaptation, would be more effective. For some healthcare students, academic achievement appears to decline when practical learning is insufficient, and this is something that must be addressed. However, this could be attributed to the sudden transition to online learning mid semester in which students did not have a chance to prepare or plan how they may need to adjust their own learning strategies. Moreover, teaching online requires a new skill set and educators may have had very little chance to upskill. It is therefore difficult to use the findings to inform future educational planning. Identifying which aspects of health education delivery are best delivered via a particular format or platform will be key to improving the efficiency of learning for organisations and accessibility of material for students. Time will tell as to the career progress of the students whose studies have been affected by COVID-19 with educators and regulators ensuring that health care professionals are supported in their learning and standards are maintained. Further research with robust methods to evaluate alternative education delivery strategies is needed to inform policy decision-making in this area.

## Data Availability

All data produced in the present work are contained in the manuscript

## Data availability statement

No data are associated with this article.

## Acknowledgements

The authors would like to thank Assim Javaid for his contribution during stakeholder meetings to guide the focus of the review and interpret findings. In addition, thanks to Professor Jane Noyes for information regarding the adaption of the GRADE approach for quantitative descriptive studies. The authors would also like to express their gratitude to Maggie Hendry and acknowledge her contribution to the Wales COVID-19 Evidence Centre report. Maggie Hendry participated in study selection, data extraction, and quality assessment.

## Extended data

Additional material one: Full search strategies http://www.primecentre.wales/resources/RR/Clean/RR00004_Supplementary_information_Healthcare_education.pdf

Additional material two Critical appraisal scores http://www.primecentre.wales/resources/RR/Clean/RR00004_Supplementary_information_Healthcare_education.pdf

Additional material three: Tool for assessing the confidence of synthesised findings from quantitative descriptive studies http://www.primecentre.wales/resources/RR/Clean/RR00004_Supplementary_information_Healthcare_education.pdf

Additional material four: Evaluation of confidence using GRADE http://www.primecentre.wales/resources/RR/Clean/RR00004_Supplementary_information_Healthcare_education.pdf

Additional material; five: excluded studies http://www.primecentre.wales/resources/RR/Clean/RR00004_Supplementary_information_Healthcare_education.pdf

## Footnotes

1 In the summer of an academic year, there are two “senior” classes (these are fourth year college students in America). The class that just graduated, known as graduating seniors, and the one that will be seniors, when fall comes around known as “oncoming senior” or “rising seniors.”

